# Seeking Help from Chatbots for Suicide Thoughts: Associations with Other Help Seeking Sources and Mental Health Symptoms

**DOI:** 10.64898/2026.08.12.26360318

**Authors:** Ruth Heo, Lauren E. McBride, Emma M. Parrish, Anthony Fulginiti, Charles T. Taylor, Colin Depp

## Abstract

**Background:** Generative AI is evolving at a rapid pace, and many individuals are utilizing chatbots for mental health support. The safety of chatbots amid suicide disclosures is a major public health focus. However, the rate and correlates of intentions to seek help from chatbots for suicide thoughts is unknown.

**Objective:** We sought to understand intentions to seek help from chatbots for suicide thoughts, compared to informal, formal, and anonymous online sources.

**Methods:** Participants with clinically significant depression or anxiety (N=58) completed the General Help Seeking Questionnaire regarding help-seeking intentions for suicide thoughts and general emotional problems. Two questions were added to assess intentions to seek help from chatbots and anonymous online sources. Wilcoxon tests were used to compare intentions to use chatbots with intentions to use anonymous online sources and with groupings of informal (e.g., friends, family) and formal (e.g., therapist, general practitioner) sources. Kendall’s correlations were used to examine correlations among groupings and individual informal and formal sources, and regression models further examined individual source associations adjusting for general help-seeking intentions. Exploratory analyses assessed whether demographic characteristics, mental health symptoms, and suicide risk were associated with help-seeking intentions for chatbots.

**Results:** Participants endorsed lower help-seeking intentions for suicide thoughts from chatbots than from informal and formal sources. Intention to use chatbots for suicide thoughts was not correlated with informal and formal sources but was correlated with anonymous online sources. At the individual source level, chatbot intentions were positively associated with intimate partners but negatively associated with outreach to friends after adjustment for general help seeking tendency. Anxiety symptom severity was positively correlated with chatbot use intentions, but not with other sources of support.

**Conclusions:** While preliminary, intentions to use chatbots for suicide thoughts appear mostly disconnected from intentions to seek help from other informal and formal supports. Future studies should evaluate the dynamics of help seeking for suicide thoughts via chatbots amidst and, perhaps in place of, other sources of support.

## Introduction

Chatbots are increasingly used for discussions about mental health. Estimates of prevalence vary widely, ranging from 5 to 45% in various samples^1–3^ and community members appear to have more positive attitudes towards using chatbots for mental health than do clinicians.^4^ A number of commentaries have suggested potential harms of using chatbots for mental health,^5,6^ such as substitution for human sources of help-seeking.^7^ There may also be potential benefits to chatbot use for mental health; for example, they may provide the opportunity to practice disclosure of mental health symptoms, which could increase confidence in discussing them with other people.^8^ Approximately 50% of individuals with thoughts of suicide do not disclose this ideation to anyone.^9,10^ Data on chatbots and suicide ideation disclosure is scarce, particularly in how intentions to use chatbots correlate with other human sources of support. In this paper, we provide preliminary data on intentions to seek help from chatbots versus formal and informal human sources for suicide thoughts.

Given the recent emergence of generative AI, data on the relationship of mental health help-seeking with chatbots compared to other sources is scarce, and most prior studies have employed convenience samples.^2,11,12^ Ueda et al.^2^ surveyed adults aged 18 to 49 about use of chatbots for mental health discussions. Within the sample, greater use of chatbots was associated with a higher likelihood of current depression and the presence of suicide ideation. When asked to rank preferences among informal and professional sources of support, chatbot users and non-users prioritized human sources of mental health support over chatbots. These findings converge with other preference-based studies, which found that while human sources of support are often preferred to chatbots, ease of access and lower cost can make chatbots attractive.^6,12^ Taken together, early data on chatbots for mental health suggest that people with mental health symptoms appear likely to use chatbots for mental health discussions, but prefer human support to chatbots.

There are several key gaps in our understanding of the correlates of chatbot use for mental health help-seeking. Prior studies have contrasted use of chatbots versus other sources in the context of general emotional problems, but did not examine use of chatbots for suicide thoughts.^2,13^ Given that suicide risk detection is a focus of safety initiatives in chatbots,^14,15^ understanding the correlates of suicide-specific help-seeking is an important gap. Moreover, studies have largely examined chatbot use in general population samples, and not in samples recruited for current mental health problems. People experiencing current mental health problems may exhibit different help seeking intentions. For example, greater depression is inversely correlated with help-seeking intentions.^16,17^ Another gap is that human sources of help-seeking (family, friends, professionals) often involve barriers such as cost, limited anonymity, and low accessibility.^6,12^ As a result, differences in help-seeking between chatbots and human sources may reflect these characteristics rather than features unique to chatbots. Including an anonymous online forum such as Reddit as a comparison condition could also help to discern whether chatbot help seeking is driven by anonymity and accessibility alone or by chatbot-specific characteristics.

In this preliminary study, we expanded a commonly used measure of mental health help-seeking intentions^18^ by adding two items assessing intentions to seek help from chatbots and anonymous online sources. We sought to understand differences in help-seeking intentions from chatbots, anonymous online sources, and both informal and formal human sources across suicide thoughts and general emotional problems. We hypothesized that 1) intentions to use chatbots for suicide thoughts would be lower than other informal and formal help seeking sources, and 2) intentions to disclose suicide thoughts to chatbots would be positively correlated with anonymous online help-seeking. We explored inter-relationships between chatbot help-seeking intentions and other individual help-seeking sources, as well as current depressive/anxious symptom severity, current suicide ideation, and demographic variables.

## Methods

### Sample

Data were derived from two ongoing longitudinal studies at UC San Diego focused on suicide help-seeking processes, and the samples were combined due to the shared focus on help-seeking. These studies have been detailed elsewhere.^19^ Participants from both studies are recruited via referrals from UC San Diego Health clinics, social media advertisements, and flyers in the community. To determine study eligibility, participants were screened for symptoms of depression using the Patient Health Questionnaire-9 (PHQ-9),^20^ symptoms of anxiety using the Generalized Anxiety Disorder-7 (GAD-7),^21^ and lifetime and current suicidal ideation and behavior using the Columbia Suicide Severity Rating Scale (C-SSRS).^22^ Participants had to have at least moderate symptoms of depression (score of 10 or higher on the PHQ-9), moderate symptoms of anxiety (score of 8 or higher on the GAD-7), or recent suicidal ideation (score of 1 or higher on the C-SSRS) to be eligible for the study.

The age range for Study 1 is 18-65 years and the age range for Study 2 is 60 years and older. The exclusion criteria for both studies are as follows: substance use disorder (excluding cannabis or tobacco) within the past three months; history of a head injury with a loss of consciousness greater than 15 minutes; presence of a neurodegenerative disorder; intellectual disability diagnosis; sensory impairments that preclude assessment (e.g., visual or auditory limitations); or if they were not able to provide informed consent based on a consent capacity assessment.^23^ Questions about help-seeking from chatbots and anonymous online sources were added to the assessment battery after data collection was underway, and this analysis includes a subset of participants who responded to questions about chatbot or anonymous online help-seeking. Both studies were reviewed by the UC San Diego Institutional Review Board, and all participants provided written informed consent to participate.

### Assessments

Analyses presented here are cross-sectional and are based on data collected via clinical interviews and self-administered questionnaires. Demographic data (e.g., age, ethnicity, race, sex) were collected by self-report. Depression and anxiety symptoms were evaluated using the Patient Health Questionnaire-9 (PHQ-9)^20^ and the Generalized Anxiety Disorder-7 (GAD-7),^21^ respectively. Current and lifetime suicide ideation, as well as past suicide attempts, were assessed via the Columbia Suicide Severity Rating Scale (C-SSRS).^22^ Participants were considered to have current suicide ideation if they endorsed having at least passive suicide ideation in the last month (ideation score of 1 or greater).

*General Help-Seeking Questionnaire:* Help-seeking intentions were assessed using the General Help-Seeking Questionnaire (GHSQ).^18^ The GHSQ has two subscales: one that assesses the participant’s likelihood to seek help for suicide thoughts and one that assesses the participant’s likelihood to seek help for emotional problems. For each subscale, participants are asked whether they would seek help from a variety of informal (i.e., partner, friend, parent, other family member) and formal (i.e., mental health professional, helpline, general doctor, religious leader) sources, from 1=extremely unlikely to 7=extremely likely. Participants answered all questions, regardless of if they had current thoughts of suicide. If participants do not have someone in their life who matches a given source, they were prompted to respond hypothetically.

We adapted this measure by adding two additional sources for each subscale: “anonymously online to people I don’t know (e.g., a Reddit forum)” and “online chatbot (e.g., ChatGPT or Gemini etc.).”

### Data Analysis

First, Wilcoxon signed-rank tests were used to compare help-seeking intentions for chatbots and anonymous online sources with help-seeking intentions for broad informal and formal source categories. Wilcoxon signed-rank tests were also used to compare source-specific help-seeking intentions across suicide thoughts and emotional problems. We then examined associations at both the broad-category and individual source levels. Kendall’s τ correlations were used to examine associations among the broad source categories and among individual sources. Regression models further examined associations between individual sources and intentions to use chatbots or anonymous online sources while adjusting for general help-seeking intentions. The general help-seeking tendency was calculated as the average of all other help-seeking items, excluding the focal predictor and outcome. Exploratory analyses examined demographic and clinical correlated of help seeking intentions for chatbots, anonymous online sources, and the broad informal and formal source categories. Due to small cell sizes, race was dichotomized for analyses (i.e., white versus non-white). Wilcoxon rank-sum tests were used for binary demographic variables and binary indicators of suicide symptoms. Kendall’s *τ* correlations were used for continuous variables, including age, depressive, and anxiety symptoms. Analyses were conducted in R version 4.5.3.^24^ Statistical significance was assessed at a two-sided a level of .05. Given the small sample size and exploratory nature of the study, findings were not adjusted for multiple comparisons.

## Results

### Sample Characteristics (Table 1)

On average, the sample was middle aged and balanced across men and women. Participants were experiencing moderately severe depressive symptoms (PHQ-9) and mild anxiety symptoms (GAD-7) (Table 1). The most common primary psychiatric diagnosis in the sample was major depressive disorder (n=47, 81%), followed by any anxiety disorder (n=5, 17%), bipolar disorder (n=2, 3%), and a psychotic disorder (n=1, 2%). Three participants (5%) did not have a primary psychiatric disorder but were eligible due to past month suicide ideation presence. A total of 86% of participants reported current suicide ideation, and 62% had a prior suicide attempt.

**Table 1.** Sample Characteristics (N=58)

|  |  | Participants |
| --- | --- | --- |
| **Age, M(SD), Range** |  | 53.1 (20.2), 18 – 82 |
| **Sex, n (%)** |  |  |
|  | Female | 34 (59%) |
|  | Male | 24 (41%) |
| **Race, n (%)** |  |  |
|  | Black or African American | 5 (9%) |
|  | White | 33 (57%) |
|  | Asian | 9 (16%) |
|  | Native Hawaiian or Other Pacific Islander | 1 (2%) |
|  | Middle Eastern or North African | 1 (2%) |
|  | More than one race | 7 (12%) |
|  | Unknown/declined to respond | 2 (3%) |
| **Ethnicity, n (%)** |  |  |
|  | Non-Hispanic or Latino origin | 50 (86%) |
|  | Hispanic or Latino origin | 8 (4%) |
| **Symptom severity** |  |  |
|  | Patient Health Questionnaire – 9, M(SD), Range | 13.0 (6.2), 1 – 26 |
|  | General Anxiety Disorder – 7, M(SD), Range | 5.1 (5.1), 0 – 18 |
|  | **CSSRS<sup>a</sup>** lifetime attempt, n (%) | 36 (62%) |
|  | **CSSRS<sup>a</sup>** current suicide ideation, n (%) | 50 (86%) |

<sup>a</sup> CSSRS=Columbia Suicide Severity Rating Scale.

### Endorsement of Chatbot Help Seeking Intentions (Table 2)

Participants endorsed lower intentions to seek help for suicide thoughts than for emotional problems across help-seeking sources (*ps* < 0.05). Help-seeking intentions for suicide thoughts were lower for chatbots compared to informal and formal sources (*M*=1.8 out of 7, SD = 1.8). In examining the distribution of endorsement of chatbot intentions, a total of 21% of the participants endorsed chatbot help-seeking intentions as 4 (neutral) or higher (1 to 7 scale) for emotional problems and 17% for suicide thoughts. Exploratory comparisons at the individual source level indicated significantly lower help-seeking intentions for chatbots than for intimate partners, friends, mental health professionals, and general doctors for both emotional problems and suicide thoughts (*ps* < .05, Supplementary Table 1).

**Table 2.** Descriptive Statistics for Help-Seeking Intentions Ratings by Source.

|  | Suicide thoughts,<br>*M (SD)* | Emotional<br>problems,<br>*M (SD)* | Wilcoxon *V* | *\|r\|* | *p* |
| --- | --- | --- | --- | --- | --- |
| Chatbot | 1.79 (1.67) | 2.14 (1.72) | 94 | 0.71 | .008** |
| Anonymous<br>online source | 1.72 (1.48) | 2.33 (1.51) | 206.5 | 0.71 | .001** |
| Informal<br>source | 3.08 (1.66) | 3.39 (1.28) | 842.5 | 0.33 | .022* |
| Formal source | 3.11 (1.39) | 3.48 (11.23) | 946 | 0.37 | .008** |

Informal sources included intimate partners, friends, parents, and other family members. Formal sources included mental health professionals, helplines, general doctors, and religious leaders. “Anyone” and “other” sources were excluded from the analysis. Wilcoxon signed-rank tests were conducted to compare help-seeking intentions across topics: general emotional problems versus suicide thoughts. *V* refers to the Wilcoxon signed-rank test statistic. Effect sizes are reported as the absolute value of *r*. Following Cohen’s thresholds for *r*, *|r|* values of $0.10 < 0.30$ , $0.30 < 0.50$ , and $^3 .50$ were interpreted as small, medium, and large effects, respectively.<sup>28,29</sup> Additional analyses were conducted to compare chatbot help-seeking intentions with formal and informal sources for general emotional problems and suicide thoughts. For general emotional problems, chatbot help-seeking intentions were lower than formal sources, mean difference = -1.45, *V* = 201, *|r|* = 0.56, $p < .001$ , and formal sources, mean difference = -1.24, *V* = 147, *|r|* = .60, $p < .001$ . For suicide thoughts, chatbot help seeking intention were lower than formal sources, mean difference = -1.45, *V* = 131.50, *|r|* = 0.62, $p < .001$ , and informal sources, mean difference = -1.31, *V* = 141, *|r|* = 0.59, $p < .001$ . \*\*\* $p < .001$ , \*\* $p < .01$ , \* $p < .05$

### Correlations with other forms of help-seeking (Figures 1 and 2)

As hypothesized, chatbot help-seeking intentions were positively correlated with anonymous online help-seeking intentions (τ = 0.25, *p* = .031) for general emotional problems (Figure 1). However, chatbot use was not significantly correlated with anonymous online help-seeking for suicide thoughts (τ = 0.17, *p* = .164). Chatbot help-seeking intentions for suicide thoughts were not correlated with intentions to seek help from informal or formal sources (informal: τ = 0.28, *p* = .194, formal: τ = 0.10, *p* = .346). In contrast, anonymous online help-seeking was positively correlated with help-seeking intentions from informal and formal sources for both suicide thoughts and general emotional problems (suicide thoughts, informal: τ = 0.28, *p* = .009; formal: τ = 0.25, *p* = .0019; general emotional problems, informal: τ = 0.28, p = .007, formal: τ =0.27, *p* = 0.01). Further exploration among individual sources (see Supplementary Figure 1) indicated that chatbot help-seeking intentions were correlated with intentions to seek help from intimate partners and helplines for suicide thoughts (*ps* < .05) and from intimate partner and anonymous online sources for general emotional problems (*ps* < .05).

**Figure 1.**
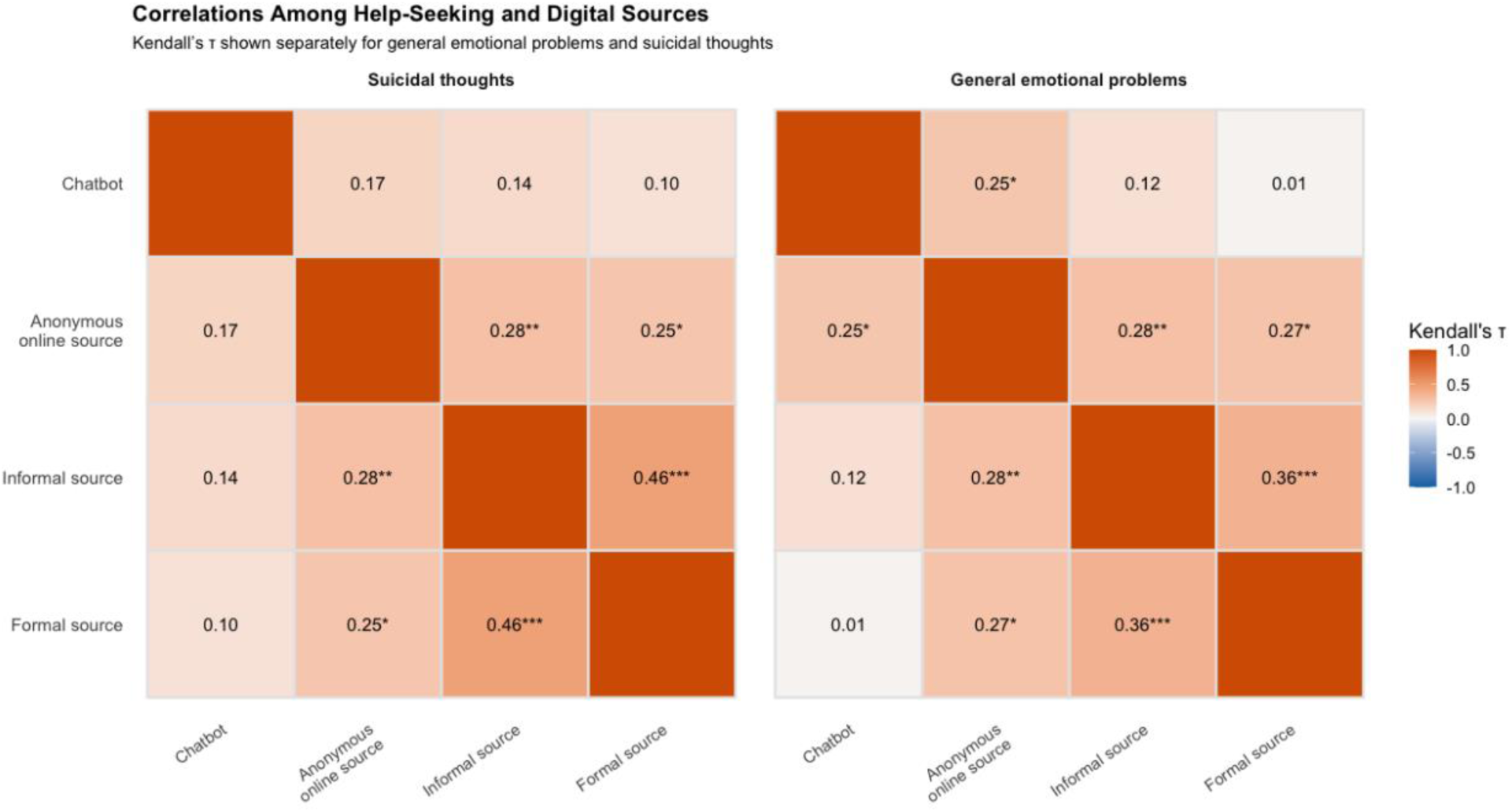
Kendall’s τ Correlations Among Aggregated Chatbot, Anonymous Online, Informal, and Formal Help-Seeking Intention Sources Informal sources included intimate partners, friends, parents, and other family members. Formal sources included mental health professionals, helpline, general doctors, and religious leaders. “Anyone” and “other” sources were excluded from the analysis. *** *p* < .001, ** *p* < .01, * *p* < .05

**Figure 2.**
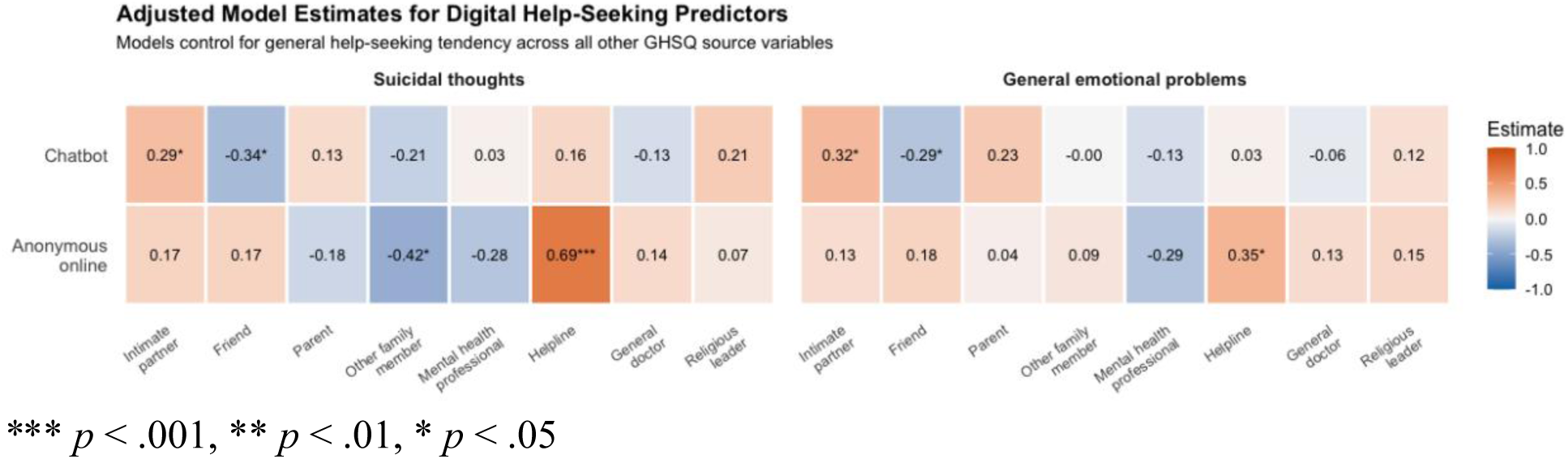
Adjusted Associations of Anonymous Online Source and Chatbot Intentions with Other Help-Seeking Sources *\**\*\* *p* < .001, ** *p* < .01, * *p* < .05

To account for generalized tendencies toward help-seeking (or negation), we adjusted for other forms of help-seeking in evaluating paired associations using regression models (see Figure 2). We found that chatbot help-seeking intentions were negatively associated with help-seeking intentions from friends for suicide thoughts (*b* = −0.34, SE = 0.14, *t* = −2.42, *p* = .019) and emotional problems (*b* = −0.29, SE = 0.14, *p* = 0.048), but remained positively associated with help-seeking intentions from intimate partners for both suicide thoughts and emotional problems (*b* = 0.29, SE = 0.14, *t* = 2.03, *p* = .047; *b* = 0.32, SE = 0.14, *t* = 2.29, *p* = .026, see Figure 2). In contrast, anonymous help seeking was positively associated with helplines for both suicide thoughts (*b* = 0.69, SE = 0.15, *t* = 4.68, *p* < .001) and emotional problems (*b* = 0.35, SE = 0.13, *t* = 2.56, *p* = .013) after adjusting for general help-seeking tendencies. Anonymous online help seeking was also negatively associated with help-seeking intentions from other family members for suicide thoughts (*b* = −0.42, SE = 0.16, *t* = −2.66, *p* = .01).

### Exploratory Associations with Demographics, Depression, Anxiety, and Suicide Ideation and Behavior

Higher chatbot help-seeking intentions were correlated with younger age for both suicide thoughts and emotional problems (τ = −0.26, *p* = .004; τ = −0.30, *p* = .016, Supplementary Table 2). A total of 31.8% of participants younger than age 50 endorsed a score of 4 (neutral) or higher for using chatbots for suicide thoughts, compared with 2.8% among participants older than age 50. No significant associations were observed between race, ethnicity, or sex and suicide help seeking intentions (Supplementary Table 2). With respect to mental health symptoms, anxiety was positively correlated with chatbot help-seeking intentions for both suicide thoughts and emotional problems (W = 0.33, *p* = .003; W = 0.31, *p* = .004, respectively). Notably, anxiety was not related to other sources of help-seeking for suicide thoughts and emotional problems. Depression was not correlated with help-seeking intentions from any sources (Supplementary Table 3). Finally, neither current suicide ideation nor lifetime suicide attempt was associated with chatbot help-seeking intentions (Supplementary Table 3).

## Discussion

In this preliminary evaluation of intentions to seek help from chatbots for suicide thoughts in a sample of people with clinically significant depression and/or anxiety (86% of whom had past month suicide ideation), we found intentions to seek help from chatbots were lower than from many informal or formal sources and lower for suicide thoughts compared to general emotional problems. On balance, there was a sizable portion of people expressing neutral or more favorable intentions to use chatbots for suicide thoughts. Notably, intentions to use chatbots for help-seeking were independent from other informal and formal help-seeking sources (although positively correlated with some individual sources, such as intimate partners and helplines). Adjusting for general help-seeking tendencies, intentions to seek help for suicide thoughts from chatbots were negatively associated with help-seeking from friends. Intentions to seek help from chatbots were correlated with anonymous online sources for general mental health issues, however, for suicide thoughts, anonymous online help-seeking intentions were related to informal and formal help-seeking, whereas chatbot help-seeking intentions were not.

Furthermore, anxiety symptom severity was positively associated with chatbot help-seeking intentions but not intentions to seek help from anonymous online sources. Pending replication, these findings suggest that help-seeking intentions for suicide thoughts with chatbots may differ from other sources of support, including anonymous online sources. Understanding user motivations for chatbot help-seeking for suicide thoughts, and its social implications such as substitution, should be a priority.

Our findings focused on suicide specific help seeking with chatbots align with prior literature on general mental health help seeking with chatbots.^2,25^ Although endorsements of intentions to use chatbots for suicide thoughts were lower than those for more general emotional problems, approximately one-third of adults under age 50 years reported neutral or more favorable intentions to use them. Chatbot help-seeking intentions for suicide thoughts were positively correlated with some sources of support, in particular intimate partners and crisis helplines. Adjusting for general intentions to seek help, chatbot help-seeking intentions were associated with lower intentions to seek help from friends. Pending replication, inverse associations between friends and chatbot help seeking intentions warrants further investigation. Since chatbots offer anonymity and ease of access, we contrasted chatbot help-seeking with anonymous online help-seeking. While correlated with chatbot help-seeking intentions, anonymous online help-seeking was associated with alternative sources of help-seeking, whereas chatbot help-seeking was not. Our findings are therefore consistent with the possibility that some individuals may view chatbots as an alternative to seeking support from friends when experiencing suicide thoughts.^26,27^ This pattern was not evident for anonymous online help-seeking, suggesting that factors beyond anonymity and accessibility may contribute to preferences for chatbot support.

In the present study, more severe anxiety was correlated with stronger intention to seek support from chatbots for suicide thoughts and emotional problems. Notably, anxiety severity was not linked to any other source of support. One hypothesis for this finding is that chatbots may provide immediate reassurance, which may be valued by persons experiencing greater anxiety.^29^ In contrast, depressive symptoms were uncorrelated with help-seeking from any source.^2,30^ Whereas Ueda et al.^2^ and Perlis et al.^27^ found that greater depressive symptoms were linked to increased chatbot use, we did not find a link between depressive symptoms and intentions to seek help from chatbots. This may be because our sample did not include participants without depressive symptoms. Although the evidence base remains limited, these findings tentatively suggest that chatbot help-seeking may not follow the pattern commonly observed in help-seeking with human sources of support, wherein more severe affective symptoms are inversely correlated with help-seeking intentions.^16,31^

This study had several limitations. Chiefly, the sample size was small, and these results need to be replicated. Relatedly, the sample size precluded more sophisticated analyses like evaluating profiles of help-seeking or moderators of effects. Data were cross-sectional and asked about intentions rather than actual help-seeking behaviors. Intentions to seek help seem to predict subsequent actual help-seeking behaviors,^18^ but it is not clear if this extends to chatbots. In respect to generalizability, our sample was skewed toward older age ranges, and given negative age associations with chatbot use, associations among help-seeking sources may have been attenuated. Many other variables would be important to consider as modifiers of chatbot help-seeking, including trust in AI, mental health stigma, digital literacy, and access to informal and formal help-seeking sources.^32^ Finally, our study did not distinguish among types of chatbots (e.g., general platforms, mental health specific, companion AI). An important direction for future research would be to understand if and how chatbot behavior may impact formal and informal human help seeking. Experimental evidence suggests agreeable or sycophantic response may increase chatbot use and reduce human social motivation, which theoretically could inhibit engagement with other sources of support.^33^

In respect to clinical implications for people considering or actively using chatbots for suicide disclosure, asking about barriers and facilitators to accessing other sources of support could be beneficial. This is especially imperative given the minimal evidence on the effectiveness of utilizing chatbots for mental health support.^34^ For future research, it would be useful to explore motivational processes that lead persons to turn toward chatbots and perhaps away from other sources of support. Potential research avenues could include understanding the lived experience of human versus chatbot help-seeking through qualitative research, as well as longitudinal research on the relationship between chatbot use and other help-seeking behaviors. Finally, a number of initiatives have begun to benchmark chatbots with respect to responses that urge outreach to professional help once suicide risk is detected.^35^ It would be essential to understand whether therapeutic redirection protocols actually lead to help-seeking outside of the chatbot. In conclusion, although a minority, there is clearly a group of people interested in using chatbots for coping with suicide thoughts and evaluating how chatbots intersect with other sources of help could inform potential safeguards for this revolutionary technology.

## Data Availability

The data are not publicly available to protect participant confidentiality.

## Acknowledgement

We are grateful to all participants for their time and participation in this study.

## Disclaimer

The views expressed in this article are those of the authors and do not necessarily reflect the position or policy of the Department of Veterans Affairs or the United States government.

## Data Availability

The data are not publicly available to protect participant confidentiality.

## Author Contribution

CD and LM contributed to the study design and drafted and revised the manuscript. RH analyzed the data and contributed to drafting and revising the manuscript. EMP, AF, and CTT reviewed the manuscript and provided feedback.

