## appendix for "Seeking Help from Chatbots for Suicide Thoughts: Associations with Other Help Seeking Sources and Mental Health Symptoms"

**Supplementary Table 1**. Paired Comparison of Chatbot Help-Seeking Intentions with Other Sources

| Source | Mean difference | Wilcoxon V | \|*r*\| | *p* |
|---|---|---|---|---|
| **Suicide Thoughts** | | | | |
| Intimate partner | −2 | 22 | | <.001*** |
| Friend | −2 | 143.5 | 0.51 | <.001*** |
| Parent | 0 | 83 | 0.19 | .161 |
| Other family member | 0 | 148.5 | 0.26 | .051 |
| Mental health professional | −3 | 37 | 0.53 | <.001*** |
| Helpline | 0 | 109 | 0.31 | .019* |
| General doctor | −1 | 150.5 | 0.49 | <.001*** |
| Religious leader | 1 | 109.5 | 0.19 | .156 |
| **General Emotional Problems** | | | | |
| Intimate partner | −2.5 | 31.5 | 0.73 | <.001*** |
| Friend | −2 | 206 | 0.52 | <.001*** |
| Parent | −0.31 | 186.5 | 0.16 | .227 |
| Other family member | 0 | 227 | 0.21 | .112 |
| Mental health professional | −3 | 90.5 | 0.72 | <.001*** |
| Helpline | 0 | 250.5 | 0.17 | .194 |
| General doctor | −2 | 161.5 | 0.59 | <.001*** |
| Religious leader | 0 | 202 | 0.22 | .102 |

Negative mean differences indicate lower intentions to seek help from chatbots than from the comparison source. Wilcoxon signed-rank tests were conducted to compare help-seeking intentions across topics: general emotional problems versus suicide thoughts. V refers to the Wilcoxon signed-rank test statistic. Effect sizes are reported as the absolute value of *r*. Following Cohen's thresholds for r, |r| values of .10 to < .30, 30 to <.50, and $^{3}$ .50 were interpreted as small, medium, and large effects, respectively.[28,29] *** $p < .001$, ** $p < .01$, * $p < .05$

**Supplementary Table 2**. Associations of Age, Sex, Race, and Ethnicity with Help-Seeking Intentions

| | Suicide Thoughts | | General Emotional Problems | |
|---|---|---|---|---|
| | Statistic τ/W | *p* | Statistic τ/W | *p* |
| **Chatbot** | | | | |
| Age [a] | -0.26 | .016* | -0.30 | .004** |
| Sex [b] | 419 | .820 | 435 | .631 |
| Race [b] | 446.5 | .200 | 399 | .049* |
| Ethnicity [b] | 187 | .699 | 140 | .123 |
| **Online** | | | | |
| Age [a] | 0.04 | .874 | -0.07 | .486 |
| Sex [b] | 371.5 | .449 | 363 | .448 |
| Race [b] | 389 | .844 | 463 | .136 |
| Ethnicity [b] | 256 | .095 | 196 | .932 |
| **Informal** | | | | |
| Age [a] | 0.03 | .740 | -0.06 | .526 |
| Sex [b] | 276.5 | .038 | 216.5 | .003** |
| Race [b] | 508 | 0.032* | 607.5 | <.0001*** |
| Ethnicity [b] | 243 | .335 | 196 | .937 |
| **Formal** | | | | |
| Age [a] | 0.329 | .832 | 0.01 | .920 |
| Sex [b] | 350 | .363 | 298 | .083 |
| Race [b] | 500 | .045* | 486.5 | .067 |
| Ethnicity [b] | 204 | .937 | 169.5 | .932 |

[a] Kendall's τ correlation. [b] Wilcoxon rank-sum tests. Wilcoxon rank-sum tests were conducted to examine between-group differences in help-seeking intentions across demographic groups. W denotes the Wilcoxon rank-sum test statistic. Sex was categorized as female or male; race as White or non-White; and ethnicity as Hispanic or non-Hispanic. *** $p < .001$, ** $p < .01$, * $p < .05$

**Supplementary Table 3.** Associations of Symptom Characteristics with Help-Seeking Intentions

| | | Suicide Thoughts | | General Emotional Problems | |
|---|---|---|---|---|---|
| | | τ/W | *p* | τ/W | *p* |
| **Chatbot** | | | | | |
| | PHQ [a] | 0.01 | .9112 | -0.02 | .865 |
| | GAD [a] | 0.33 | .003** | 0.31 | .004* |
| | Lifetime suicide attempt [b] | 361.5 | .455 | 349.5 | .397 |
| | Past-month suicide ideation [b] | 148 | .112 | 162 | .331 |
| **Anonymous online** | | | | | |
| | PHQ [a] | 0.09 | .398 | 0.06 | .585 |
| | GAD [a] | 0.06 | .603 | 0.09 | .392 |
| | Lifetime suicide attempt [b] | 329 | .155 | 316.5 | .172 |
| | Past-month suicide ideation/behavior [b] | 200.5 | 1 | 196.5 | .942 |
| **Informal** | | | | | |
| | PHQ [a] | -0.11 | .234 | -0.14 | .154 |
| | GAD [a] | 0.06 | .603 | 0.08 | .395 |
| | Lifetime suicide attempt [b] | 370 | .681 | 366 | 0.635 |
| | Past-month suicide ideation/behavior [b] | 265.5 | .141 | 240.5 | .365 |
| **Formal** | | | | | |
| | PHQ [a] | 0.01 | .935 | 0.05 | .571 |
| | GAD [a] | 0.06 | .515 | 0.15 | .133 |
| | Lifetime suicide attempt [b] | 358 | .547 | 315.5 | 0.199 |
| | Past-month suicide ideation/behavior [b] | 213.5 | .769 | 168 | .476 |

[a] = Kendall tau [b] = Wilcoxon rank-sum test. Wilcoxon rank-sum tests were conducted to examine between-group differences in help-seeking intentions across CSSRS groups. W denotes the Wilcoxon rank-sum test statistic. *** $p < .001$, ** $p < .01$, * $p < .05$

**Supplementary Figure 1**. Correlations among Individual Help-seeking Sources

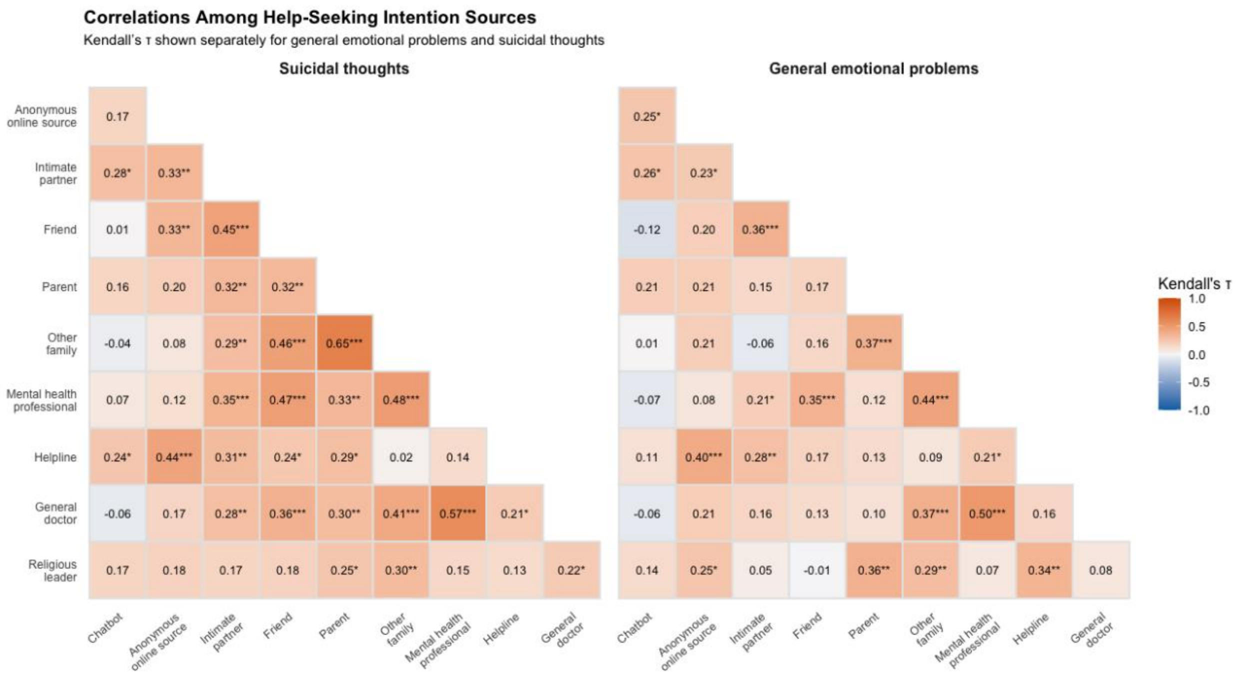


*** $p < .001$, ** $p < .01$, * $p < .05$
